# Assessing the association between contraceptive agency and preference-aligned fertility management among Ugandan women: A 12-month prospective cohort study

**DOI:** 10.64898/2026.08.27.26361582

**Authors:** Birabwa Catherine, Wasswa Ronald, Amongin Dinah, Ghosh Rakesh, Phillips Beth, Challa Sneha, Gomez Rouselinne, Atuyambe Lynn, Liu Jenny, Waiswa Peter, Holt Kelsey

## Abstract

**Background:** There has been a proliferation of new person-centered and human rights-based contraception measures in recent years, though their application in research remains limited. Improved measures offer an opportunity to examine how contraceptive decision-making agency relates to individuals’ ability to act in line with their contraceptive preferences. We sought to assess the association between contraceptive agency and subsequent Preference-aligned Fertility Management (PFM) over 12 months in a cohort of women in rural Uganda.

**Methods:** We analyzed data from a prospective cohort study conducted in five largely rural Ugandan districts from 2022 to 2024. Data were collected at baseline, 6 and 12 months from a convenience sample of women who were new users of contraception or not using contraception. We used mixed-effects logistic regression models to examine the association between baseline Agency in Contraceptive Decisions Scale overall and subscale scores and future PFM Index scores at 6 and 12 months, assessing whether associations varied over time using interaction terms for follow-up time point. We used interactions between agency scores and follow-up visit to assess whether associations differed between the 6- and 12-month visits. We assessed effect modification by age group and baseline contraceptive method category using three-way interaction terms and predicted probabilities.

**Results:** The analytic sample comprised 2,227 women. The percentage of women practicing PFM increased from 85.7% at baseline to 93.3% at 12 months. A one-unit increase in Agency in Contraceptive Decisions Scale score was associated with higher odds of subsequent PFM (aOR: 1.68, 95% CI: 1.10– 2.54). Subscales 3 (knowledge aligned with preferences) and 4 (control over use or non-use) of the Agency in Contraceptive Decisions Scale were significantly associated with future PFM (aOR: 1.31, 95% CI: 1.04–1.66 and aOR: 1.27, 95% CI: 1.06–1.51, respectively). The association between overall contraceptive agency and PFM did not differ between the 6- and 12-month visits. Three-way interaction tests suggested that the associations between the overall Agency in Contraceptive Decisions Scale score and the PFM outcomes varied jointly by age group and baseline contraceptive method category: overall PFM Index (p<0.001), PFM1 (p=0.011), and PFM2 (p<0.001).

**Conclusion:** Our findings suggest that higher levels of contraceptive agency may help women act in line with their contraceptive preferences. Increasing women’s knowledge and control over contraceptive use may be particularly essential for preferred contraceptive use. The findings also suggest that the association between contraceptive agency and PFM may vary by women’s age group and the method of choice, though further exploration is necessary to examine this influence.

## Background

Ensuring that women and couples can freely make informed decisions about whether and when to use contraception is a fundamental component of reproductive rights [1]. This can help women and couples achieve their reproductive goals in ways that align with their preferences and lived realities, with potential benefits of overall improved quality of life and well-being [2]. However, evidence from sub-Saharan Africa (SSA) shows low decision-making autonomy regarding modern contraceptive use among partnered women [3]. In Uganda, the literature indicates that fewer than 40% of women using contraception, including adolescent girls, are actively involved in decisions regarding contraceptive use [4,5]. This implies that such women may be using or not using contraception against their desires, yet such discrepancies have not been extensively measured or monitored. Conventional contraception outcome measures, including (modern) contraceptive prevalence and unmet need, focus on uptake or use, and inadequately capture the extent to which an individual’s contraception preferences are fulfilled [6]. This gap may mask issues of reproductive coercion or systemic failure to meet individual contraception needs [5].

More recently, person-centered and human rights-based alternatives to conventional contraception measures have been published [6,7]. However, their application in research and practice remains limited. The Preference-aligned Fertility Management (PFM) Index is a person-centered, rights-based measure that captures the alignment between desired and actual contraceptive use or non-use, including whether users’ current methods are desired. The PFM Index was validated among in Nigeria and Uganda and has also been applied among Nigerian adolescents [8–10]. The measure places emphasis on meeting individuals’ self-defined needs, without assuming use is always a positive outcome. PFM Indicator 1 assesses concordance between desired and actual contraceptive use (PFM 1) and Indicator 2 measures whether users’ specific methods are desired (PFM 2); the two are combined in the PFM Index. Other studies have explored the performance of similar metrics, including ‘contraceptive concordance’ [11] and ‘non-preferred and misaligned contraceptive method use’ [12,13]. Despite growing interest in measuring alignment between individuals’ contraceptive preferences and behaviors, longitudinal studies examining predictors of preference-use alignment over time have not been conducted.

The present study focuses on individual contraceptive agency as a potential determinant of PFM. Agency has been associated with contraceptive *use* in low- and middle-income countries [14–16], though its relationship with new person-centered contraception measures such as the PFM Index has not been explored. Contraceptive agency refers to an individual’s ability to make and act on their decisions regarding pregnancy prevention, whatever those decisions may be [17–19]. An individual with contraceptive agency is aware of their right to choose, has the self-efficacy and control to make decisions aligned with their values, has access to accurate and appropriate information, and critically reflects on possible constraints on their choices [17,19]. The new Agency in Contraceptive Decisions Scale, developed and validated in Nigeria and Uganda and comprised of four subscales, presents an opportunity to better measure contraceptive agency and explore its relationship with subsequent PFM [9,20]. Uganda is a pertinent location to explore the relationship between agency and PFM given that, similar to other SSA countries [21,22], Uganda faces multiple persistent challenges that may limit individuals’ ability to act in line with their preferences, including low levels of women’s empowerment [23].

This study aimed to assess the association between Agency in Contraceptive Decisions Scale scores and future PFM Index scores among women aged 15-45 years in Uganda, and to examine whether this relationship is modified by the contraceptive method chosen and the woman’s age. We analysed survey data that were collected during the Innovations for Choice and Autonomy project at three time points (T1=baseline, T2=6 months, T3=12 months). The findings contribute to the existing body of knowledge on person-centered and rights-based contraception measures.

## Methods

### Conceptual framework

This study draws on existing literature to conceptualise the hypothesised relationship between contraceptive agency and future PFM [8,9,17,20] – **Figure 1**. Several individual and interpersonal factors were considered as potential confounders of the relationship, including women’s age, economic status, education level, and marital status; the degree to which they reported discussing contraception with peers and/or partners; and their baseline PFM. We considered one’s chosen contraceptive method and their age as potential effect modifiers, based on a hypothesis that adolescents face unique social and structural barriers to having their contraceptive needs met that may stymie them regardless of their decision-making agency in a way that is less relevant than for older women. It was also hypothesised that the characteristics of the chosen contraceptive method, such as associated side effects, frequency and mode of administration, and privacy of use, may influence an individual’s ability to use a preferred method freely, and modify the effect of one’s agency on their ability to act in line with their preferences. We acknowledge that other factors at community or macro levels, including access to quality services and social norms, can impact an individual’s agency and PFM [9,17], but these were not assessed in the present study.

**Figure 1.**
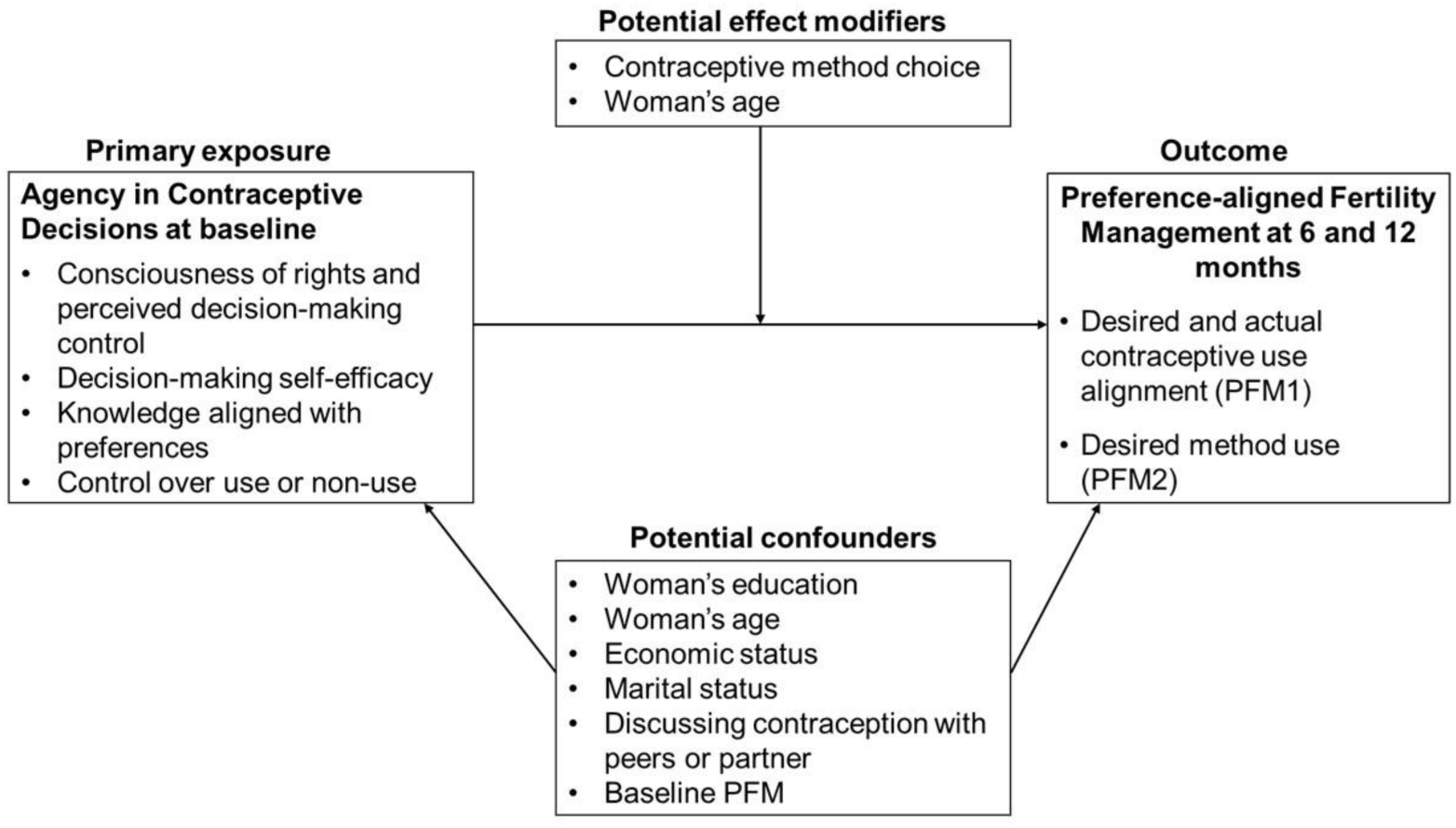
Hypothesized relationship between contraceptive agency and preference-aligned fertility management over time.

### Data source and study population

We used data from the Innovations for Choice and Autonomy (ICAN) prospective cohort study conducted in five Ugandan districts: Oyam, Kole, and Lira in Lango subregion of Northern Uganda, and Mayuge and Iganga in the Busoga subregion of Eastern Uganda. More than 80% of people living in the two subregions resided in rural areas, and contraceptive use prevalence among women in union was 40.9% in Busoga and 37.7% in Lango [24]. The ICAN Uganda cohort enrolled 2,422 sexually active women aged 15-45 years at baseline. Eligible participants included new contraceptive users, defined as women who had started using any contraceptive within two weeks before the baseline survey, as well as those who were not using any contraception or doing anything to prevent pregnancy. Data were collected between November 2022 and May 2024, including baseline, 6- and 12-month follow-up, by 10 female research assistants, fluent in English and respective local languages (Langi in Lango, Lusoga in Busoga). Participants provided written informed consent before each interview. Our analysis included 2,227 participants who were interviewed at all three data collection points **(Figure 2**). The study received ethical approval from Makerere University School of Public Health Research and Ethics Committee (SPH-2022-212), Uganda National Council for Science and Technology (HS1087ES), and University of California, San Francisco Institutional Review Board (21-34470).

**Figure 2:**
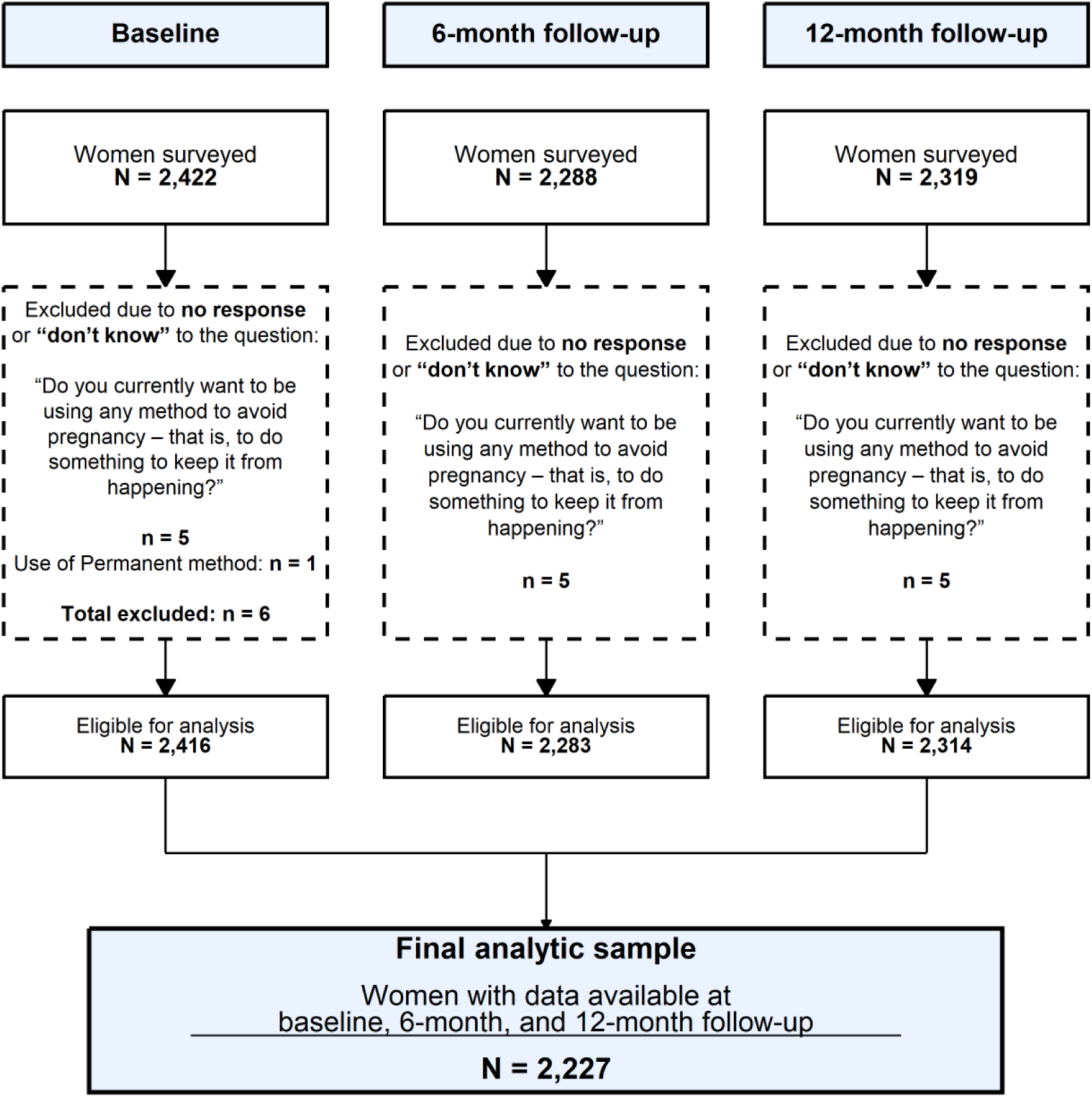
Sample size derivation.

### Study variables

#### Outcomes

PFM was measured using the validated PFM Index [9]. The dichotomous index (1=PFM, 0=no PFM) is derived from three questions that jointly measure the extent to which an individual’s contraceptive use behavior aligns with their contraceptive preferences. The index comprises two indicators: indicator1, which captures alignment between an individual’s desired and actual contraceptive use (hereafter PFM1) and indicator 2, which assesses alignment between the method(s) currently used and method-specific preferences (hereafter PFM2). Detailed conceptualization, scoring procedures, and validation of the PFM Index are described by Holt et al [9]. Our analysis included the composite PFM Index, PFM1, and PFM2.

#### Exposure

Contraceptive agency at baseline was measured using the validated Agency in Contraceptive Decisions Scale. This scale measures an individual’s ability to make and act on their contraceptive decisions, assessed using 15 items, with each item scored on a four-point Likert scale, ranging from 0=Strongly no to 3=Strongly yes. The overall agency score was computed as the mean of all item responses, with higher scores indicating greater agency in contraceptive decision -making. The scale comprises four subscales: consciousness of rights and perceived decision-making control (Subscale1), decision-making self-efficacy (Subscale2), knowledge aligned with preferences (Subscale3) and control over use or non-use (Subscale4). Additional details on the Agency in Contraceptive Decisions Scale are described by Challa et al [20].

#### Baseline covariates

We measured key sociodemographic and economic factors hypothesized to influence contraceptive agency and ability to act on preferences (Figure 1). These included: woman’s age (15-19 years, 20-24 years, 25+ years), highest education level (none or primary school vs secondary or higher), and socioeconomic status (SES). Socioeconomic status was derived using the EquityTool, a validated instrument designed to assess relative household wealth, asset ownership and housing characteristics such as dwelling conditions, access to financial services, and household assets [25]. Each item was weighted according to the EquityTool’s standard Uganda-specific scoring algorithm, and the resulting scores were aggregated to generate a composite wealth index. Women were subsequently grouped into three SES tertiles: low/poor (1), middle (2), and high/rich (3) households. Other covariates included marital status (married or unmarried) and frequency of discussion about contraceptives. The latter was derived from two survey items that asked how often women discussed pregnancy prevention with (1) women in their families and (2) peers outside their families. Each item was rated on a four-point ordinal scale: 1=Never, 2=Yearly, 3=Monthly, and 4=Weekly. Responses to the two items were averaged to obtain a composite measure reflecting the overall frequency of discussion, which was then further categorized into three ordered levels: never/rarely (≤1.5), occasionally (>1.5–≤2.5), and frequently (>2.5). These cut-off points corresponded to midpoints between adjacent ordinal categories, preserving the scale’s inherent order. Contraceptive use at baseline was also incorporated for effect modification analyses as a categorical variable, distinguishing between non-users, users of short-acting reversible contraceptives (SARCs) and users of long-acting reversible contraceptives (LARCs). The SARCs included injectables, pills, condoms, lactational amenorrhea, and fertility awareness methods, while LARCs included implants and intra -uterine devices.

### Data analysis

We used descriptive statistics to summarize baseline participant characteristics. Categorical variables were presented as frequencies and percentages, while continuous variables were summarized using means and standard deviations.

The association between baseline contraceptive agency and future PFM was examined using mixed - effects logistic regression models. Separate models were fitted for the overall agency in contraceptive decisions scale score and each of its four subscale scores each PFM outcome: PFM Index, PFM 1, and PFM 2. Each model included follow-up visit as a categorical fixed effect and random intercepts for geographic cluster and participant. Because several sub-counties had few participants, we combined them into 12 geographic clusters within the five study districts based on geographic proximity and similar socioeconomic characteristics. The participant-level random intercept accounted for repeated observations within each woman, while the cluster-level random intercept accounted for correlation among women within the same geographic cluster. For woman i in geographic cluster j at follow-up visit t, the model was specified as:

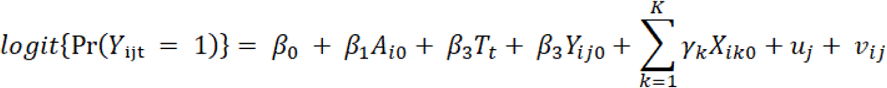

where Yᵢⱼₜ denotes the PFM outcome for woman i in cluster j at follow-up visit t, where t represents either the 6- or 12-month follow-up visit. Aᵢ₀ denotes the overall baseline contraceptive agency score or one of the four baseline subscale scores; Tₜ denotes follow-up visit; Yᵢⱼ₀ denotes the corresponding PFM outcome at baseline; and Xᵢₖ₀ denotes baseline covariate k for woman i. The terms uⱼ and vᵢⱼ denote the random intercepts for geographic cluster and participant, respectively.

To assess whether the association between baseline contraceptive agency and future PFM differed at 6 and 12 months follow-up visits, we fitted separate models that included an interaction between each baseline contraceptive agency score and follow-up visit. The model was specified as:

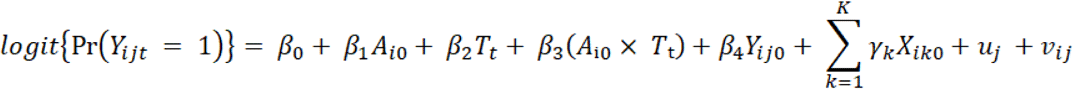

Where β₃ is the coefficient for the interaction between baseline contraceptive agency and follow-up visit. A statistically significant interaction was interpreted as evidence that the association between baseline contraceptive agency score and PFM differed between the 6- and 12-month visits.

We then assessed whether women’s age and baseline contraceptive method category jointly modified the association between baseline contraceptive agency and subsequent PFM. For each agency measure and PFM outcome, we fitted the following model:

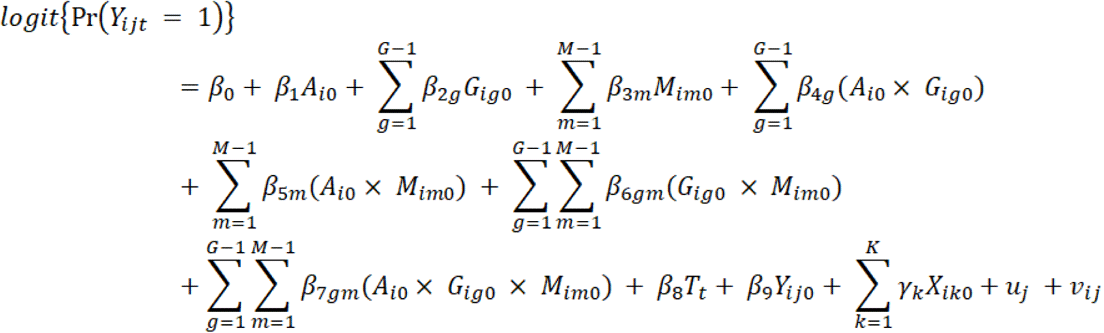

Here, Gᵢg₀ denotes indicator g for baseline age group, and Mᵢm₀ denotes indicator m for baseline contraceptive method category. The coefficients β₇gm represent the three-way interactions between baseline contraceptive agency, age group, and baseline contraceptive method category. Joint Wald tests were used to assess the statistical significance of the three-way interaction terms. Adjusted predicted probabilities were estimated across the range of agency scores for each combination of age group and baseline contraceptive method category to illustrate the interaction patterns. All statistical analyses were conducted using Stata version 18, and R version 4.5.0 was used for data visualization. Statistical significance was assessed at p<0.05.

## Results

### Participant characteristics

More than half of the participants were aged 25–45 years (55.5%), nearly three-quarters had no formal education or had completed primary education (73.7%), and almost all were married or had a partner (97.5%) - **Table 1**. Approximately six in ten women were using long-acting reversible contraceptives at baseline (58.9%), while 24.7% were using short-acting reversible contraceptives and 16.3% were not using a contraceptive method. The mean overall Agency in Contraceptive Decisions score was 2.6 (SD = 0.4), with mean subscale scores ranging from 2.4 (SD = 0.7–0.8) to 2.7 (SD = 0.4).

**Table 1:** Baseline participant characteristics (N=2227)

| *Categorical variables* | *Frequency* | *Percentage* |
| --- | --- | --- |
| **Women's age group** |  |  |
| 15-19 | 271 | 12.2 |
| 20-24 | 721 | 32.4 |
| 25-45 | 1,235 | 55.4 |
| **Women's education level** |  |  |
| None or Primary school | 1,641 | 73.7 |
| Secondary/Higher | 586 | 26.3 |
| **Socioeconomic status tertiles** |  |  |
| Low | 804 | 36.1 |
| Middle | 745 | 33.5 |
| High | 678 | 30.4 |
| **Marital status** |  |  |
| Unmarried | 55 | 2.5 |
| Married/Has a partner | 2,172 | 97.5 |
| **Frequency of discussions on pregnancy prevention** |  |  |
| Never/Rarely | 227 | 10.2 |
| Occasionally | 699 | 31.4 |
| Frequently | 1,301 | 58.4 |
| **Contraceptive method category** |  |  |
| Non-user | 364 | 16.3 |
| SARCs | 551 | 24.7 |
| LARCs | 1,312 | 58.9 |
| **Continuous variables** |  |  |
|  | **Mean** | **Standard deviation** |
| **Agency in Contraceptive Decisions Scale scores** |  |  |
| Overall score | 2.6 | 0.4 |
| Subscale 1 (*Consciousness of rights and perceived decision-making control*) | 2.7 | 0.4 |
| Subscale 2 (*Decision-making self-efficacy*) | 2.6 | 0.6 |
| Subscale 3 (*Knowledge aligned with preferences*) | 2.4 | 0.7 |
| Subscale 4 (*Control over use or non-use*) | 2.4 | 0.8 |

SARCs = short-acting reversible contraceptives; LARCs = long-acting reversible contraceptives. Agency in Contraceptive Decisions Scale scores range from 0 to 3, with higher scores indicating greater contraceptive agency.

### Preference-aligned Fertility Management over a one-year period

The percentages of women meeting the overall PFM, PFM1, and PFM2 criteria increased over the 12- month follow-up period. Overall PFM increased from 85.7% at baseline to 89.9% at 6 months and 93.3% at 12 months. PFM1 increased from 86.1% to 93.8% and 94.0%, while PFM2 increased from 86.4% to 90.7% and 93.5% at the corresponding visits (**Figure 3**).

**Figure 3:**
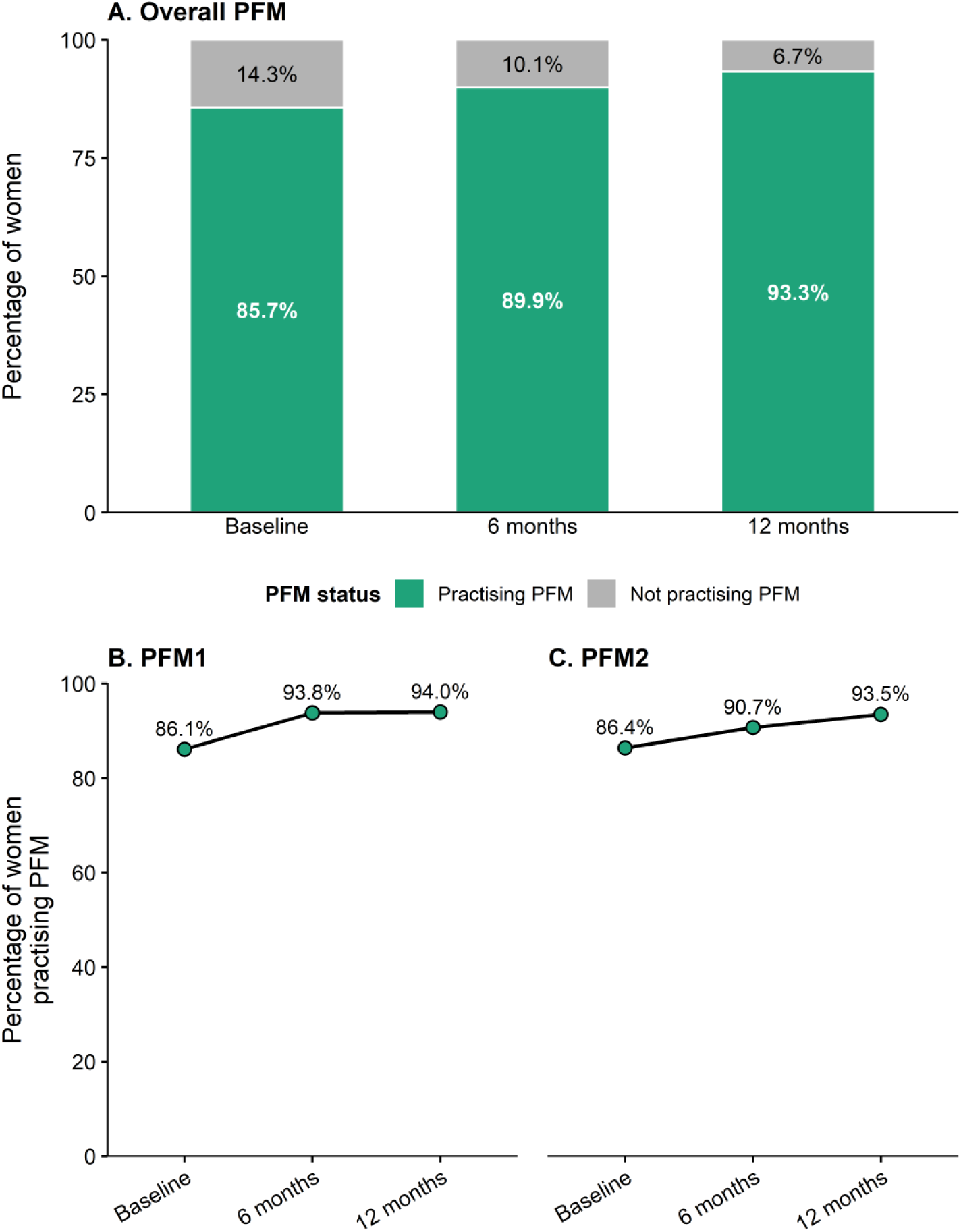
Preference-aligned Fertility Management over 12 months (N=2,227). *Note: PFM = preference-aligned fertility management. PFM1 assesses alignment between desired and actual contraceptive use or non-use. PFM2 assesses alignment between the contraceptive method or methods currently used and method-specific preferences*.

### Association between baseline contraceptive agency and future preference-aligned fertility management

Overall, higher baseline contraceptive agency was associated with greater odds of future PFM, although the magnitude of association varied across agency subscales and PFM outcomes (**Table 2**). A one-unit increase in the overall Agency in Contraceptive Decisions Scale score was associated with 68% higher odds of overall PFM (aOR = 1.68, 95% CI: 1.10–2.54), and more than two-fold higher odds of PFM1 (aOR = 2.13, 95% CI: 1.39–3.26).

**Table 2:** Adjusted association between contraceptive agency and future preference-aligned fertility management (N=2227)

| Agency in Contraceptive Decisions Scale baseline score | Future PFM Index score<br>aOR (95% CI) | Future PFM 1 score<br>aOR (95% CI) | Future PFM 2 score<br>aOR (95% CI) |
| --- | --- | --- | --- |
| **Overall composite score** | 1.68 (1.10-2.54)* | 2.13 (1.39-3.26)*** | 1.49 (0.98-2.25) |
| **Subscale 1:**<br>Beliefs about rights and decision-making control | 1.34 (0.97-1.85) | 1.44 (1.04-2.00)* | 1.24 (0.88-1.74) |
| **Subscale 2:**<br>Decision-making self-efficacy | 0.98 (0.74-1.29) | 1.01 (0.70-1.46) | 0.90 (0.69-1.16) |
| **Subscale 3:**<br>Knowledge aligned with preferences | 1.31 (1.04-1.66)* | 1.63 (1.31-2.03)*** | 1.23 (0.99-1.53) |
| **Subscale 4:**<br>Control over use or non-use | 1.27 (1.06-1.51)** | 1.33 (1.16-1.52)*** | 1.28 (1.02-1.61)* |

aOR = adjusted odds ratio; CI = confidence interval; PFM = preference-aligned fertility management. Estimates are from separate repeated-measures mixed-effects logistic regression models fitted for each contraceptive agency score and PFM outcome, using observations from the 6- and 12-month follow-up visits. Models included random intercepts for geographic cluster and participant identifier to account for clustering and within-participant correlation, and were adjusted for baseline PFM, age group, education level, marital status, socioeconomic status, and discussion on pregnancy prevention. \*p < 0.05, \*\*p < 0.01, \*\*\*p < 0.001

Among the agency subscales, control over use or non-use (Subscale 4) showed the most consistent positive association across all three PFM outcomes. A one-unit increase in Subscale 4 was associated with higher odds of overall PFM (aOR = 1.27, 95% CI: 1.06–1.51), PFM1 (aOR = 1.33, 95% CI: 1.16–1.52), and PFM2 (aOR = 1.28, 95% CI: 1.02–1.61). Knowledge aligned with preferences (Subscale 3) was associated with higher odds of overall PFM (aOR = 1.31, 95% CI: 1.04–1.66) and PFM1 (aOR = 1.63, 95% CI: 1.31–2.03). Beliefs about rights and decision-making control (Subscale 1) was positively associated with PFM1 only (aOR = 1.44, 95% CI: 1.04–2.00), while decision-making self-efficacy (Subscale 2) was not significantly associated with any PFM outcome.

Associations between baseline contraceptive agency and future PFM were generally consistent across the 6- and 12-month follow-up periods (**Supplementary Table S1**). We found no evidence that the strength of associations differed by follow-up time for the overall PFM or PFM2 outcomes (all interaction p-values > 0.05). For PFM1, the association was greater at the 6-month time point than at 12 months for the overall Agency in Contraceptive Decisions Scale score (interaction p = 0.002) and Subscale 1 (beliefs about rights and decision-making control) (interaction p = 0.049).

### Effect modification by contraceptive method and woman’s age

**Table 3** shows evidence that the association between contraceptive agency and PFM varied jointly by age group and contraceptive method category. The Wald test for the three-way interaction was statistically significant across all three outcomes: overall PFM (p<0.001), PFM 1 (p=0.011), and PFM 2 (p<0.001). For the agency subscales, Subscale 1, reflecting rights and decision-making, showed evidence of a three-way interaction for the overall PFM (p=0.036) and PFM 1 (p=0.002), but not for PFM 2. Subscale 2, reflecting decision-making self-efficacy, showed evidence of interaction only for PFM 2 (p=0.034). Subscale 3, reflecting knowledge aligned with preferences, showed evidence of interaction for PFM 1 (p=0.014) and PFM 2 (p=0.018). There was no evidence of a three-way interaction for Subscale 4, reflecting control over use or non-use, across any of the three outcomes. **Figure 4** presents predicted probabilities of overall PFM Index scores by baseline overall contraceptive agency score, age group, and contraceptive method category. Subscale-specific predicted probabilities are presented in the **supplementary**.

**Figure 4:**
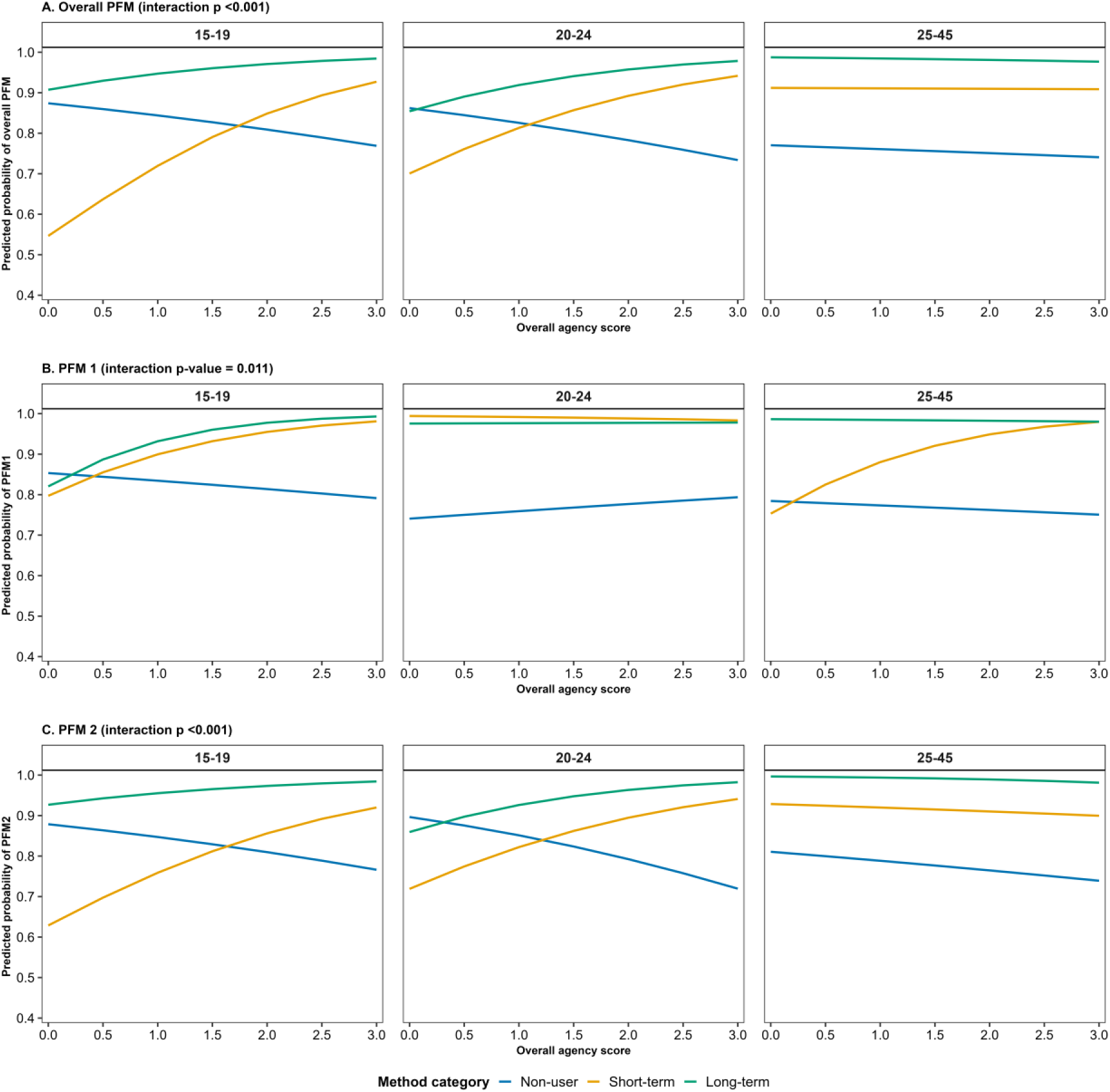
Predicted probability of preference-aligned fertility management by overall contraceptive agency score and subgroups defined by age and contraceptive use at baseline. *Predicted probabilities were estimated from mixed-effects logistic regression models that included a three-way interaction between baseline contraceptive agency score, age group, and baseline contraceptive method category. Models adjusted for follow-up visit, the corresponding PFM outcome at baseline, education level, marital status, socioeconomic status tertile, and frequency of discussion about pregnancy prevention, with random intercepts for geographic cluster and participant. The p-value reported for each PFM outcome is from the joint Wald test of the contraceptive agency score × age group × baseline contraceptive method category interaction and assesses whether the association between contraceptive agency and PFM varied jointly by age group and baseline contraceptive method category*.

**Table 3:** P-values from three-way interaction terms examining differences in the relationship between baseline contraceptive agency and future PFM by age group and contraceptive method category.

| Agency in contraceptive decisions scale score | PFM overall<br>p-value | PFM 1<br>p-value | PFM 2<br>p-value |
| --- | --- | --- | --- |
| Overall contraceptive agency score | 0.000 | 0.011 | 0.000 |
| Subscale 1: Rights and decision-making | 0.036 | 0.002 | 0.462 |
| Subscale 2: Decision-making self-efficacy | 0.238 | 0.226 | 0.034 |
| Subscale 3: Knowledge aligned with preferences | 0.084 | 0.014 | 0.018 |
| Subscale 4: Control over use or non-use | 0.330 | 0.527 | 0.525 |

*P-values are from Wald tests of the three-way interaction between contraceptive agency score, age group, and baseline contraceptive method category. Models adjusted for baseline PFM, education level, marital status, socioeconomic status tertile, and discussion on pregnancy prevention, with random intercepts for geographic cluster and participant identifier.*

## Discussion

We assessed the relationship between contraceptive agency (measured by the Agency in Contraceptive Decisions Scale) and preference-aligned contraceptive use (measured by the PFM Index) over a one-year period in a cohort of over 2,000 women of reproductive age from rural Uganda who were almost all married. The findings showed that baseline agency was significantly associated with future PFM, after accounting for baseline PFM levels and other potential confounders. Agency in Contraceptive Decisions subscales 3 (knowledge aligned with preferences) and 4 (control over use or non-use) showed the most consistent associations with PFM. Additionally, interaction models and predicted probabilities suggested that the association between contraceptive agency and PFM varied jointly by age and baseline contraceptive method category.

Our finding of an association between contraceptive agency and PFM aligns with evidence that agency and empowerment are associated with traditional measures of contraceptive use that focus on uptake without considering women’s preferences [14–16]. Continued research in this area across geographies will be important to characterize the degree to which new measures produce results that differ from standard contraceptive use indicators. Regardless, however, using person-centered outcome measures such as PFM remains an imperative as the international family planning field continues to move towards full operationalization of long-standing human rights-based standards that demand a focus on women’s preferences.

Our finding that subscales 3 and 4 of the Agency in Contraceptive Decisions Scale were most consistently positively associated with the PFM Index and its indicators suggests that having accurate knowledge aligned with one’s preferences (subscale 3) and control over contraceptive use or non-use (subscale 4) may play particularly important roles in shaping women’s ability to practice PFM compared to their awareness about reproductive rights and perceived control (subscale 1) or decision -making self-efficacy (subscale 2). Nonetheless, subscale 1 showed a significant positive relationship with PFM1, suggesting that consciousness of rights and perceived control, as components of agency, may play a particularly crucial role in shaping the alignment between desired and actual contraceptive use. These findings highlight the need to strengthen informational/knowledge empowerment of women and to address issues concerning contraceptive coercion.

Effect modification analyses suggested that the association between contraceptive agency and PFM varied jointly by women’s age and baseline contraceptive method category. These patterns may reflect differences in social influences, including norms and partner support, and access to quality contraceptive services, which may shape women’s ability to exercise agency in contraceptive decisions [17]. While these subgroup patterns were based on interaction models and predicted probabilities, further research is needed to confirm these patterns and understand the mechanisms underlying them.

### Strengths and limitations

Our study advances the understanding and informs future applications of two human rights-based family planning measures. Nonetheless, the study has the following limitations: First, the observational design means we cannot rule out unmeasured confounding, and thus our results cannot be interpreted causally. Second, because we used convenience sampling, the findings may not be generalisable to all women in the study districts. Notably, our sample consists largely of married women in rural settings, and future research with unmarried women and in urban and humanitarian settings is warranted.

## Conclusion

Our results suggest that increased contraceptive agency may increase married women’s ability to use or not use contraception in line with their preferences in rural Uganda and that the role of agency may be particularly salient for younger contraceptive users. The findings suggest the importance of empowering women with awareness of their right to choose and the self-efficacy and control to make contraceptive decisions, based on their values, with preference-aligned information and support, while critically reflecting on possible constraints to their choices. Future studies should further examine this association using rigorous methods for analyzing observational data and exploring the relationship in other settings and with different populations, such as women living in urban or humanitarian settings and unmarried women.

## Author contributions

**BC**: conceptualisation, investigation, methodology, formal analysis, writing – original draft preparation, writing – review and editing. **WR**: conceptualisation, investigation, methodology, formal analysis, data curation, visualisation, writing – original draft preparation, writing – review and editing. **AD**: conceptualisation, investigation, funding acquisition, writing – review and editing. **GRakesh**: methodology, formal analysis, visualisation, writing – review and editing. **PB**: conceptualisation, writing – review and editing. **CS**: data curation, writing – review and editing. **GRouselline**: data curation, writing – review and editing. **AL**: conceptualisation, methodology, funding acquisition, writing – review and editing. **LJ**: conceptualisation, funding acquisition, writing – review and editing. **WP**: conceptualisation, funding acquisition, methodology, writing – review and editing. **HK**: conceptualisation, formal analysis, visualisation, funding acquisition, writing – review and editing. All authors reviewed and approved the final version of the manuscript.

## Funding

This work was supported by the Gates Foundation, INV-009958. The Gates Foundation did not play a role in the design and conduct of the study as well as the analysis.

## Conflict of interest

The authors declare no conflicts

## Data availability statement

The data supporting the results in this manuscript will be made available in an online repository.

## Supplementary materials

**Supplementary Table S1:** Association between contraceptive agency scores and PFM outcomes at 6- and 12-month follow-up visits. Estimates are adjusted odds ratios (aORs) from mixed-effects logistic regression models including an interaction between agency score and survey wave. Models were adjusted for baseline PFM, age group, education level, marital status, socioeconomic status, and discussion on pregnancy prevention, and included random intercepts for cluster and participant identifier. Interaction p-values assess whether associations differed between 6- and 12-month follow-up visits.

**Outcome: Overall PFM**
| Agency scale | Interaction<br>p-value | 6-month<br>aOR (95% CI) | 12-month<br>aOR (95% CI) |
| --- | --- | --- | --- |
| Overall composite score | 0.5787 | 1.61 (1.11-2.34) | 1.83 (1.02-3.27) |
| Subscale 1: Rights & decision-making | 0.6424 | 1.41 (1.11-1.80) | 1.25 (0.69-2.26) |
| Subscale 2: Self-efficacy | 0.5426 | 0.94 (0.71-1.25) | 1.03 (0.73-1.47) |
| Subscale 3: Knowledge aligned with<br>preferences | 0.4658 | 1.24 (0.93-1.66) | 1.44 (1.05-1.97) |
| Subscale 4: Control over use/non-use | 0.5782 | 1.24 (1.05-1.46) | 1.32 (1.01-1.71) |

**Outcome: PFM1**
| Agency scale | Interaction p-value | 6-month aOR (95% CI) | 12-month aOR (95% CI) |
| --- | --- | --- | --- |
| Overall composite score | 0.0013 | 2.73 (1.80-4.15) | 1.63 (0.97-2.72) |
| Subscale 1: Rights & decision-making | 0.0772 | 1.83 (1.33-2.53) | 1.10 (0.63-1.92) |
| Subscale 2: Self-efficacy | 0.2948 | 1.10 (0.76-1.60) | 0.92 (0.59-1.43) |
| Subscale 3: Knowledge aligned with preferences | 0.2288 | 1.81 (1.52-2.16) | 1.45 (1.00-2.11) |
| Subscale 4: Control over use/non-use | 0.9844 | 1.33 (1.18-1.51) | 1.33 (0.96-1.83) |

**Outcome: PFM2**
| Agency scale | Interaction p-value | 6-month aOR (95% CI) | 12-month aOR (95% CI) |
| --- | --- | --- | --- |
| Overall composite score | 0.4470 | 1.38 (0.94-2.01) | 1.67 (0.93-3.01) |
| Subscale 1: Rights & decision-making | 0.8275 | 1.27 (1.01-1.62) | 1.20 (0.66-2.20) |
| Subscale 2: Self-efficacy | 0.3862 | 0.84 (0.65-1.09) | 0.97 (0.69-1.37) |
| Subscale 3: Knowledge aligned with preferences | 0.4966 | 1.16 (0.87-1.56) | 1.34 (0.99-1.82) |
| Subscale 4: Control over use/non-use | 0.3471 | 1.24 (0.99-1.55) | 1.34 (1.02-1.76) |

**Supplementary Figure S1:**
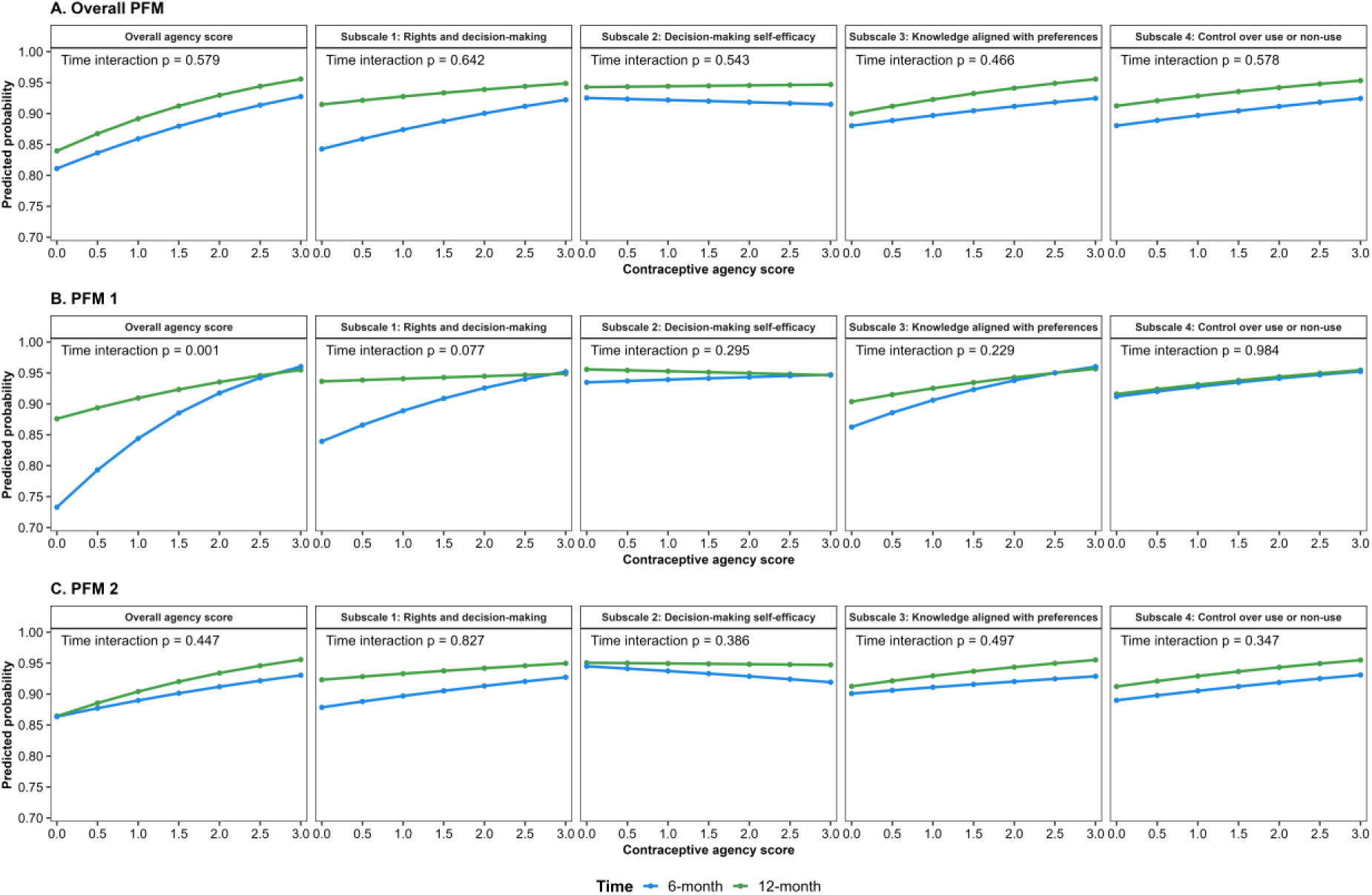
Predicted probabilities of preference-aligned fertility management by baseline contraceptive agency score and follow-up time.

Predicted probabilities of preference-aligned fertility management at the 6- and 12-month follow-up visits according to baseline contraceptive agency scores. Curves are based on adjusted mixed-effects logistic regression models including an interaction between contraceptive agency score and follow-up time. Models were adjusted for baseline PFM status, age group, education, marital status, socioeconomic status, and frequency of discussion on pregnancy prevention, with random intercepts for cluster and participant. The p-value shown in each panel corresponds to the contraceptive agency score # follow-up time interaction.

**Supplementary Figures S2–S5.** Predicted probabilities of preference-aligned fertility management by contraceptive agency subscale, age group and contraceptive method category.

The figures show predicted probabilities for the four Agency in Contraceptive Decisions Scale subscales: rights and decision-making, decision-making self-efficacy, knowledge aligned with preferences, and control over use or non-use.

The subscale-specific figures show that the interaction patterns were not uniform across agency dimensions. Consistent with Table 3, there was evidence of three-way interaction for Subscale 1 with the overall PFM and PFM 1, for Subscale 2 with PFM 2, and for Subscale 3 with PFM 1 and PFM 2. Subscale 4 showed no evidence of interaction across the three PFM outcomes.

**Figure S2.**
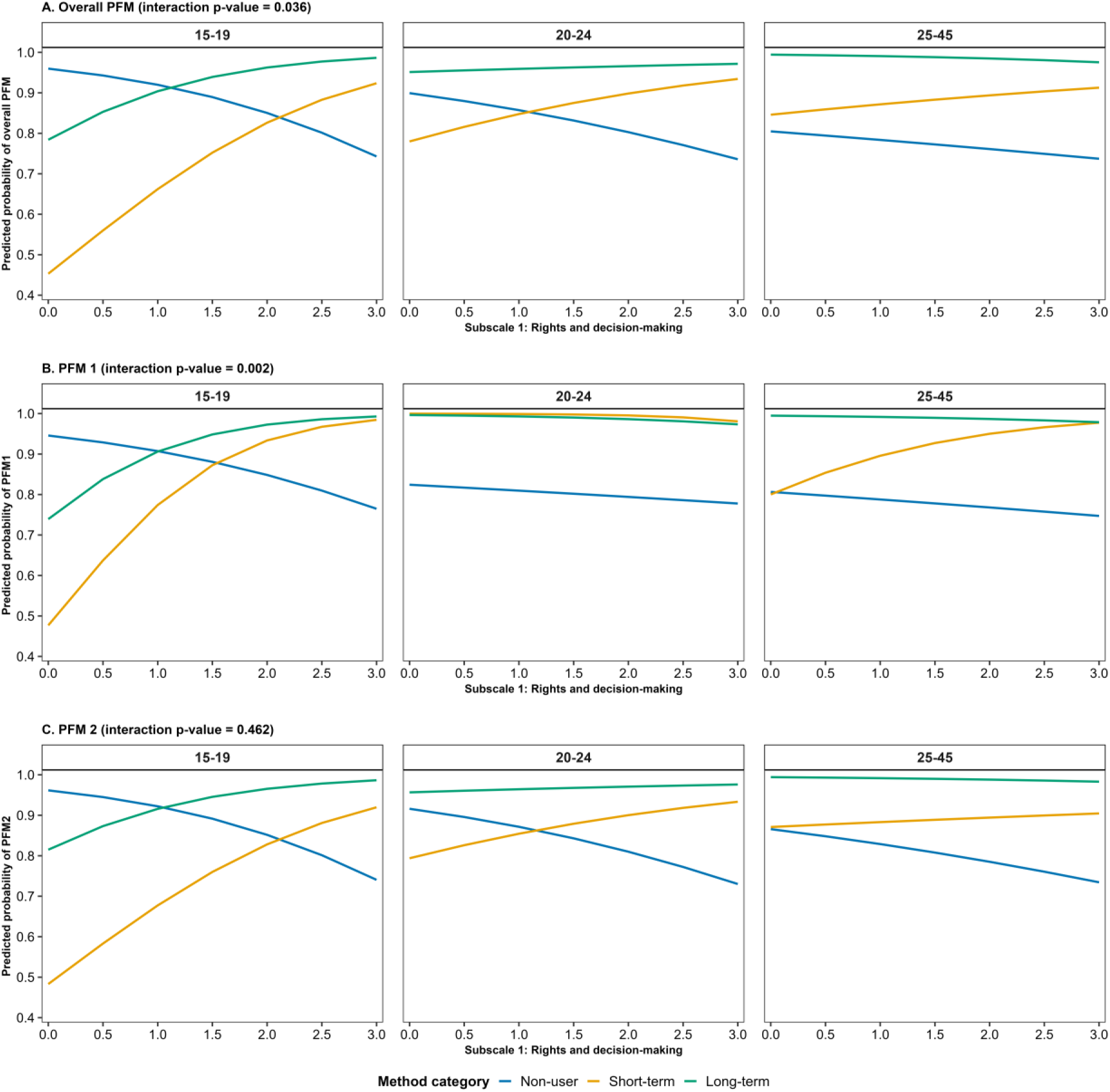

**Figure S3.**
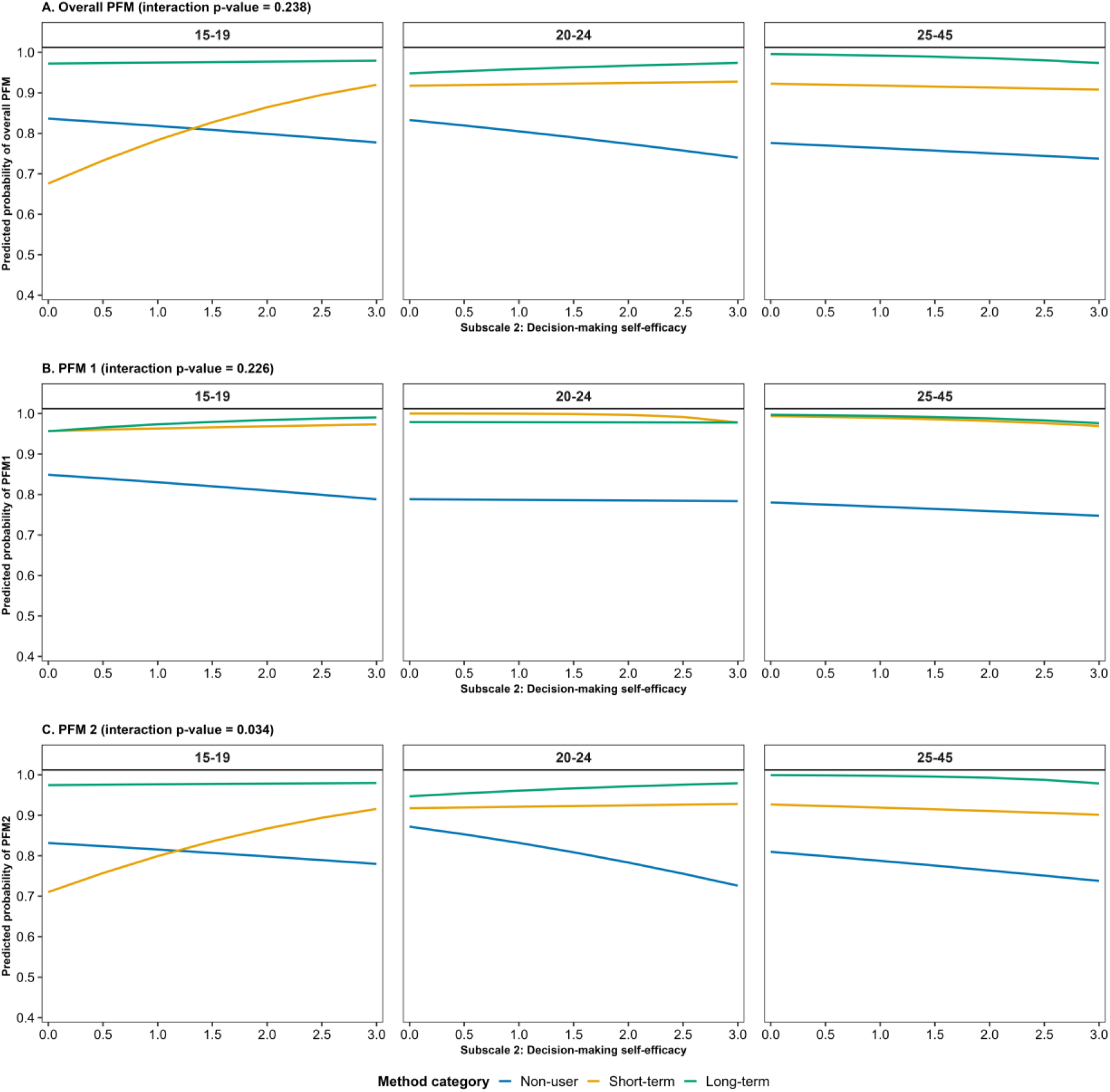

**Figure S4.**
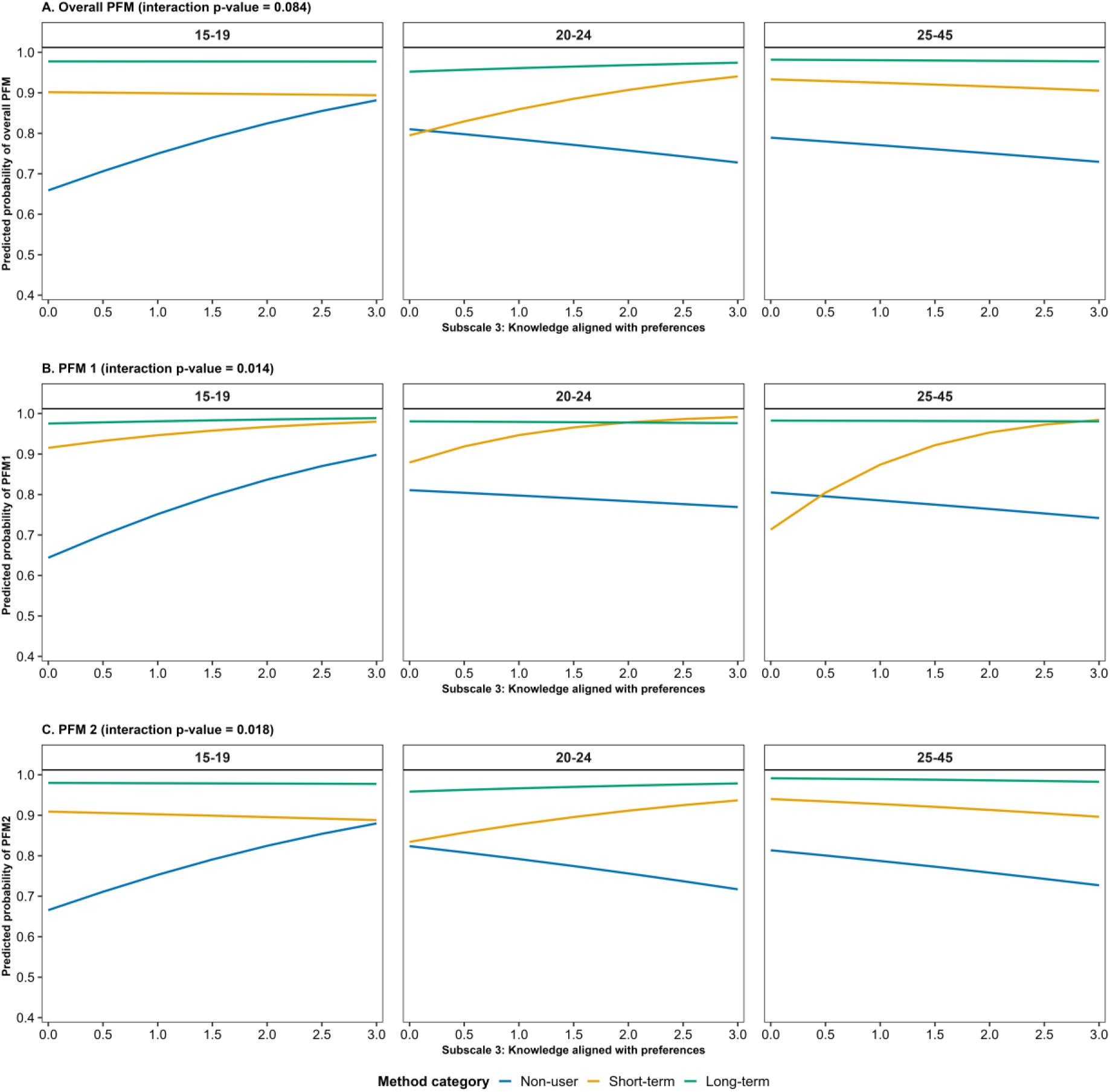

**Figure S5.**
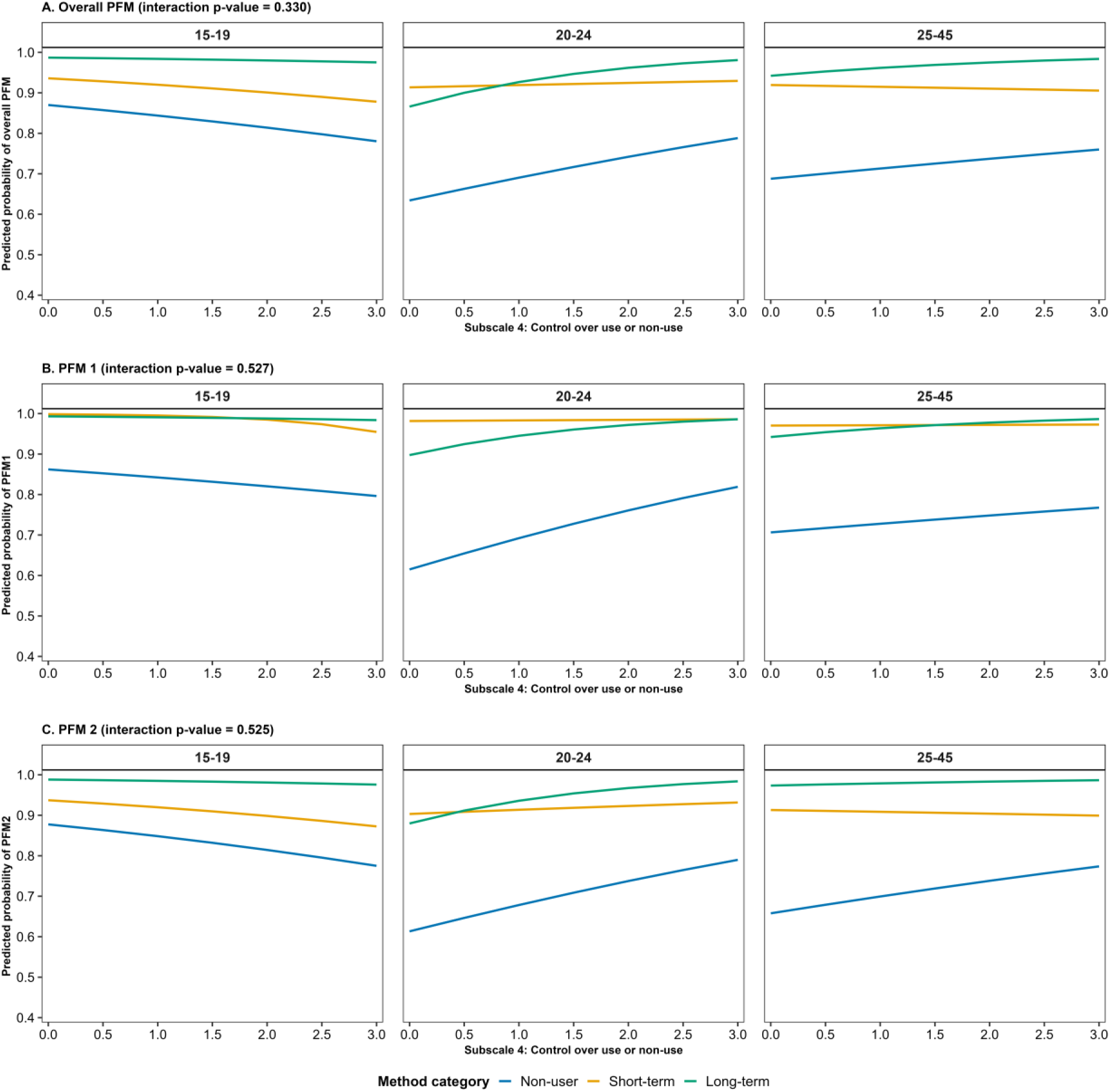

## Notes

### Competing Interest Statement

The authors have declared no competing interest.

### Author Declarations

The study received ethical approval from Makerere University School of Public Health Research and Ethics Committee (SPH-2022-212), Uganda National Council for Science and Technology (HS1087ES), and University of California, San Francisco Institutional Review Board (21-34470).

